## Supplementary for "Comparative evaluation of plasma biomarkers of *Schistosoma haematobium* infection in endemic populations from Burkina Faso"

**SUPPLEMENTARY INFORMATION**

**Supplementary Figure 1. Venn proportional diagram showing the comparison of CAA and cfDNA testing results (DeepVenn, https://arxiv.org/abs/2210.04597).**

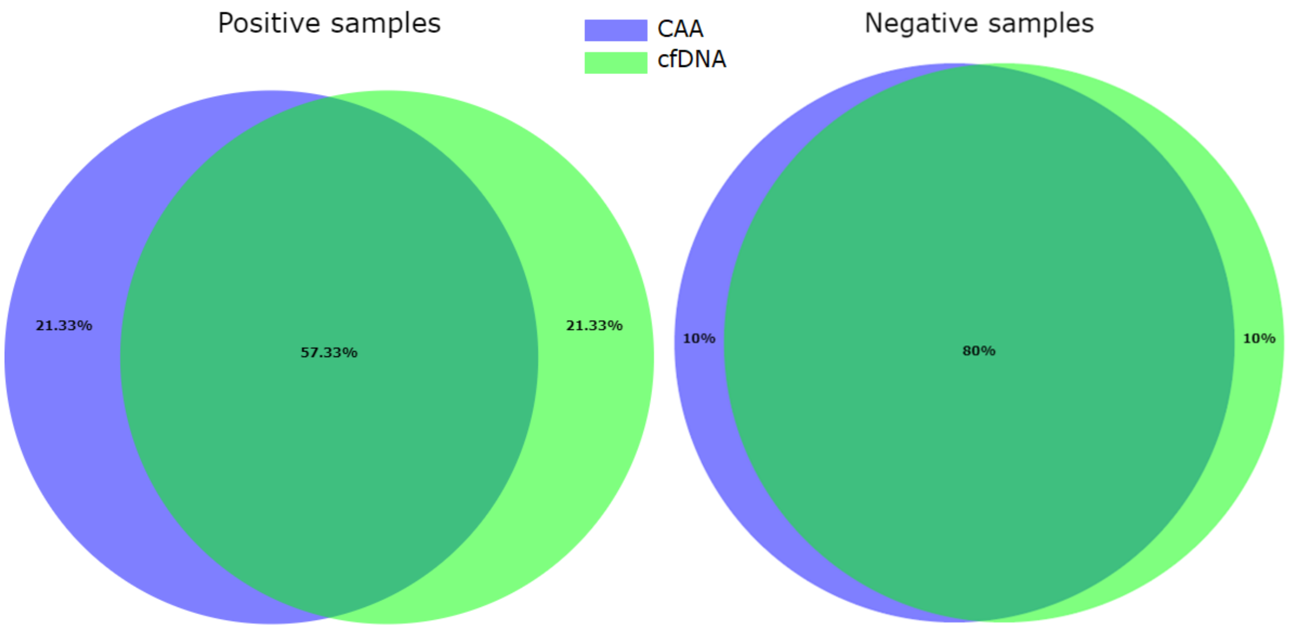

**Supplementary Figure 2. ROC curves illustrating the diagnostic performance of anti-*S.haematobium* antibodies compared to Circulating Anodic Antigen.**

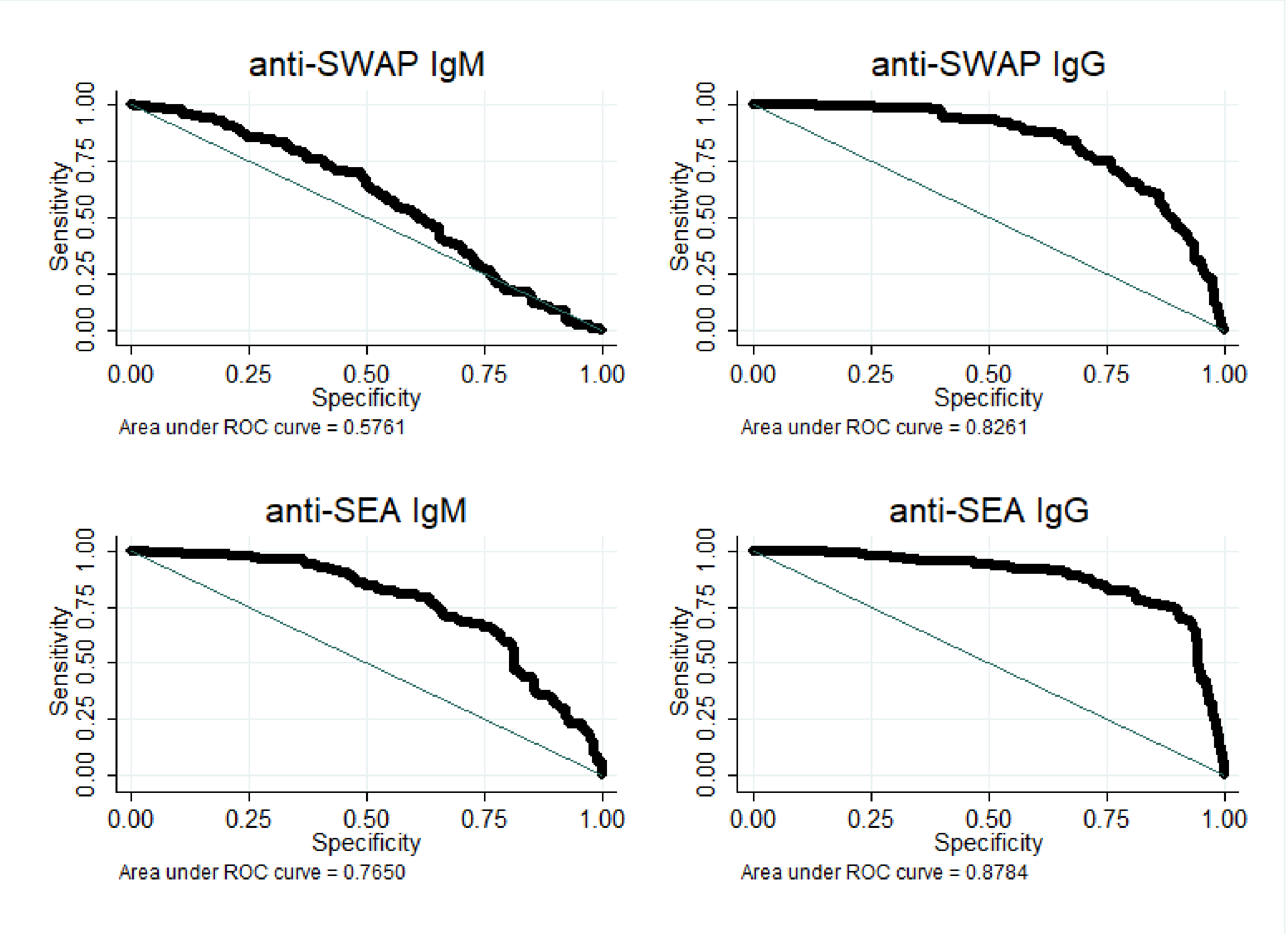

**Supplementary Figure 3. ROC curves illustrating the diagnostic performance of anti-*S.haematobium* antibodies compared to Composite Reference Standard.**

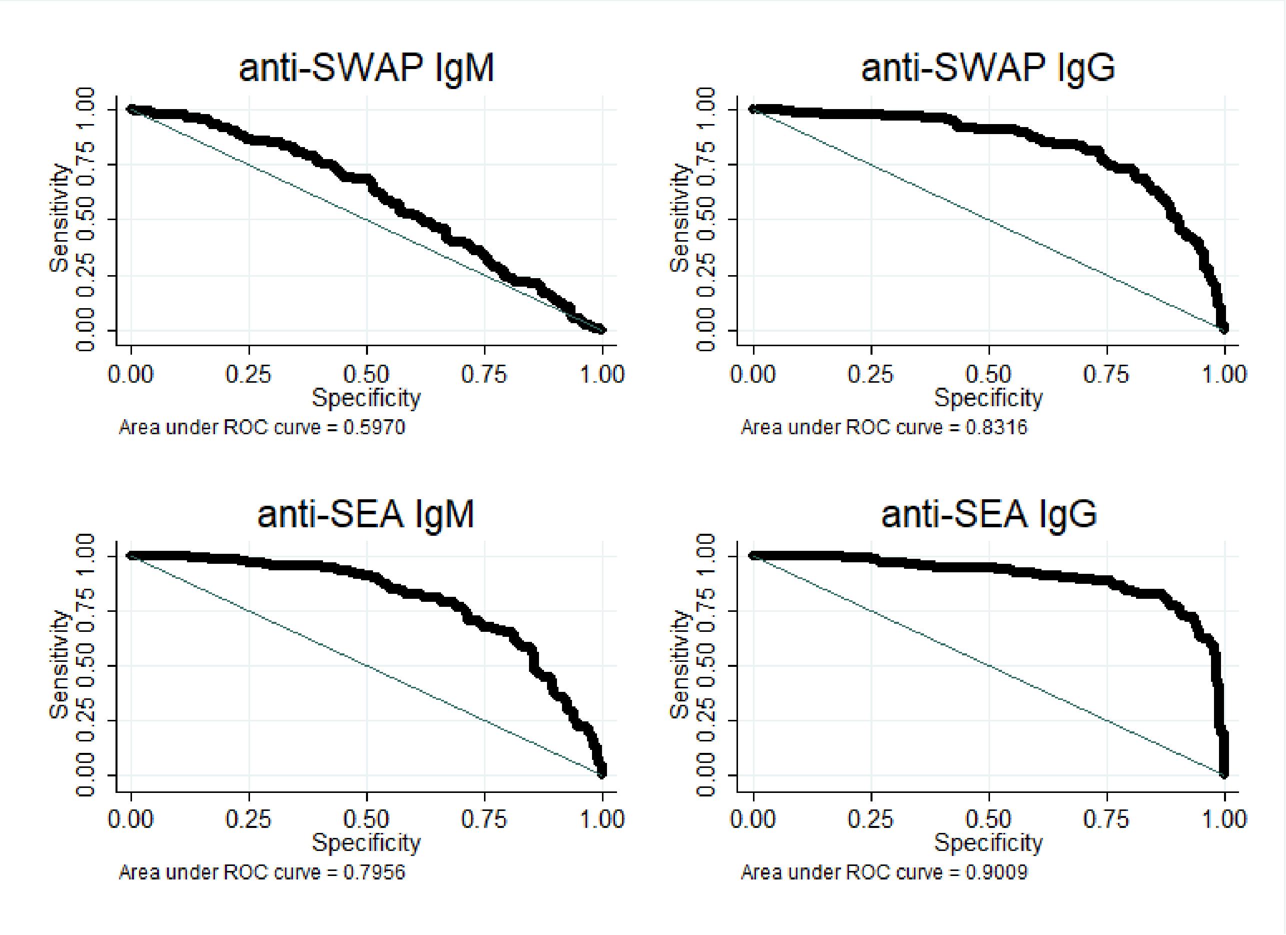

**Supplementary Figure 4. ROC curves illustrating the diagnostic performance of anti-*S.haematobium* antibodies compared to Latent Class Analysis.**

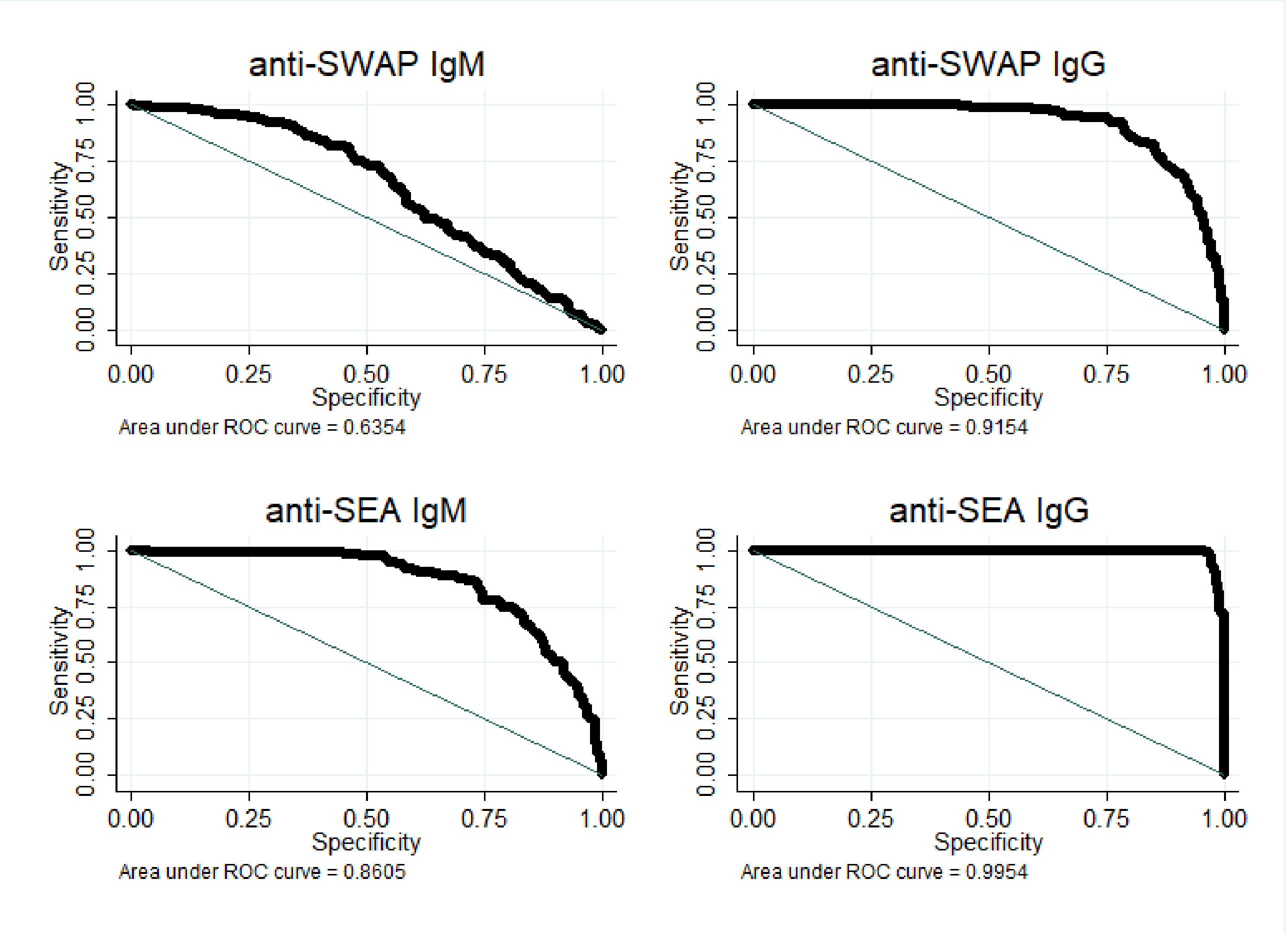
